# Shared genetic and environmental influences between the broad avoidant/restrictive food intake disorder phenotype and neurodevelopmental traits: a twin study

**DOI:** 10.64898/2026.06.19.26356081

**Authors:** Baiyu Qi, Liv Hog, Paul Lichtenstein, Sebastian Lundström, Henrik Larsson, Cynthia M. Bulik, Ralf Kuja-Halkola, Mark J. Taylor, Lisa Dinkler

## Abstract

**Importance:** Avoidant/restrictive food intake disorder (ARFID) is a feeding and eating disorder characterized by extremely restricted dietary variety and/or quantity resulting in significant physical health impairment and psychosocial dysfunction. ARFID frequently co-occurs with neurodevelopmental conditions, yet the extent to which this co-occurrence reflects shared genetic or environmental influences remains largely unknown, as few twin or genetic studies of ARFID have been conducted.

**Objective:** To examine the extent to which genetic and environmental influences contribute to the association between a broad ARFID phenotype and neurodevelopmental traits.

**Design, Setting, and Participants:** Population-based twin study using data from the Child and Adolescent Twin Study in Sweden, including 30,374 twins born 1992–2008.

**Main Outcomes and Measures:** A broad ARFID phenotype was identified using a composite measure derived from parent reports and national health registers between ages 6 and 12 years. Parents completed measures of neurodevelopmental traits at age 9 or 12 years, including autism (subdomains: social communication problems and restricted/repetitive behaviors), attention-deficit/hyperactivity disorder (ADHD, subdomains: inattention and impulsivity/hyperactivity), tic disorders, learning disorders, oppositional defiant disorder, conduct disorder, obsessive–compulsive disorder (OCD), sensory perception problems, and sleep problems. Phenotypic associations were estimated using polyserial correlations. Bivariate twin models decomposed variance and covariance into genetic and environmental components.

**Results:** Phenotypic correlations with the broad ARFID phenotype ranged from 0.18 (95% CI: 0.15-0.21) for OCD to 0.36 (95% CI: 0.33-0.38) for autism. Broad genetic correlations (rH; additive plus dominant genetic influences) ranged from 0.27 (95% CI: 0.21-0.33) for conduct disorder to 0.52 (95% CI: 0.44-0.60) for autism–restricted/repetitive behaviors. Genetic factors explained 77% to 95% of all phenotypic correlations. Non-shared environmental correlations were minimal to small, with the largest observed for autism (0.17; 95% CI: 0.08-0.26).

**Conclusions and Relevance:** The broad ARFID phenotype shares substantial genetic influences with a number of neurodevelopmental traits. These findings suggest that the frequent co-occurrence of ARFID with neurodevelopmental traits largely reflects shared genetic influences rather than overlapping environmental influences, supporting the conceptualization of ARFID within a broader neurodevelopmental framework.

## Introduction

Avoidant/restrictive food intake disorder (ARFID) is a feeding and eating disorder characterized by persistent restriction in the amount and/or variety of food intake that is not driven by concerns about body weight or shape, leading to significant impairments in weight, growth, nutrition, and psychosocial functioning.^1–3^ First formally recognized in the Diagnostic and Statistical Manual, 5th Edition (DSM-5) in 2013,^3^ ARFID can occur across the lifespan but is most commonly identified in childhood and early adolescence.^4^ ARFID frequently co-occurs with neurodevelopmental conditions. A meta-analysis of 21 studies reported an autism prevalence of 16.3% (range: 2–88%) among individuals with ARFID, and an ARFID prevalence of 11.4% (range: 2–28%) among individuals with autism; however variation between studies is substantial.^5^ Similarly, children with attention-deficit/hyperactivity disorder (ADHD) have demonstrated higher levels of ARFID-related picky eating and sensory processing difficulties, with selective eating positively associated with ADHD symptom severity.^6^ ARFID has also been associated with moderate to severe obsessive-compulsive disorder (OCD) symptoms in adults,^7,8^ although this association was weaker in pediatric samples.^8^ Two recent population-based studies using the same cohort as the current study reported elevated risks of multiple neurodevelopmental conditions among children with ARFID, including autism (prevalence in ARFID, 12.1-13.8%), ADHD (16.9-17.5%), intellectual disability (9.1%), learning disorders (2.1-9.4%), oppositional defiant disorder (ODD; 19.4%), OCD (11.0%), conduct disorder (1.5-5.9%), and tic disorders (2.3-14.8%), with the highest odds ratios for autism (9.68-13.71) and intellectual disability (10.27).^9,10^ Together, these findings indicate high prevalences of multiple neurodevelopmental conditions among individuals with ARFID. However, little is known about whether these associations reflect shared genetic or environmental mechanisms.

Importantly, both ARFID and many neurodevelopmental conditions are highly heritable. In a recent population-based twin study, the heritability of ARFID was estimated at 70-85%.^11^ Similarly, substantial genetic influences have been reported for neurodevelopmental conditions, such as autism (64%-91%),^12^ ADHD (74%),^13^ learning disorders (40-70%),^14^ tic disorders (30%-40%),^15^ ODD (34%–73%),^16^ conduct disorder (40-50%),^17^ and OCD (50%).^18^ Molecular genetic studies suggest some genetic overlap between ARFID and neurodevelopmental conditions. One study found that ARFID score was nominally positively associated with polygenic risk scores for autism, but not ADHD.^19^ Another study estimated SNP-based genetic correlations of broadly and clinically defined ARFID with autism, ADHD, and OCD, and reported a significant genetic correlation only between clinically defined ARFID and ADHD (r_g_ = 0.21, SE = 0.10).^20^ However, these studies were limited to molecular genetic overlap captured by common variants with a small number of traits and did not quantify the relative contributions of genetic and environmental factors to phenotypic associations between ARFID and neurodevelopmental traits.

To address these gaps, the current study used a large population-based twin cohort to determine the extent to which associations between a broad ARFID phenotype and neurodevelopmental traits reflect shared genetic and environmental influences. Understanding these shared etiological pathways is important for clarifying the developmental origins of ARFID, identifying at-risk individuals, and informing detection and intervention strategies for ARFID. Notably, while prior research has focused on categorical diagnoses, neurodevelopmental conditions are increasingly conceptualized as continuously distributed traits, which are genetically linked to corresponding clinical diagnoses.^21^ Examining continuous traits may therefore provide a more sensitive proxy of underlying liability than categorical diagnoses alone.

## Methods

### Participants

Data came from the Child and Adolescent Twin Study in Sweden (CATSS), a nationwide longitudinal study of twins born in Sweden since July 1, 1992.^22^ The study includes a wide range of psychiatric and neurodevelopmental phenotypes and is linked to national population health registers. The present study included twins born between 1992 and 2008 who participated in the CATSS baseline assessment at ages 9 or 12 years (response rate ∼69%). Zygosity of same-sex twins was determined using a validated panel of genetic markers for most twin pairs or by a validated questionnaire assessing twin similarity when genetic data were unavailable.^23,24^ Only twin pairs with a high probability (>95%) of correct classification were included. The CATSS study was approved by the Regional Ethical Review Board in Stockholm, Sweden. Informed consent was obtained from parents. For the current study, we excluded twins with unknown zygosity or missing co-twin, resulting in a final sample of 30,374 individuals (15,187 complete pairs). Three prior studies using overlapping CATSS data described phenotypic associations between the broad ARFID phenotype and neurodevelopmental traits and estimated its heritability.^9–11^ The present study addresses a distinct question by quantifying the extent to which associations between the broad ARFID phenotype and neurodevelopmental traits are attributable to shared genetic and environmental influences.

### Measurements

#### Assessment of ARFID

A broad ARFID phenotype was identified using a previously developed composite measure based on information relevant to DSM-5 criteria from multiple data sources, including parent reports, diagnosis and procedure data from the National Patient Register, and medication data from the Prescribed Drug Register.^11^ To be classified as having ARFID, children were required to meet DSM-5 Criterion A (avoidant or restrictive eating associated with clinically significant consequences, such as low weight, nutritional deficiency, dependence on nutritional supplements, or psychosocial impairment) and Criterion C (eating disturbance not attributable to anorexia nervosa, bulimia nervosa, or concerns about weight or shape) between ages 6 and 12 years. Information to evaluate Criterion B (lack of available food or culturally sanctioned practices) was not available. Individuals were not excluded based on Criterion D, as it is difficult to determine in an epidemiological context whether the eating disturbance is attributable to another medical or psychiatric condition.

#### Assessment of neurodevelopmental traits

Neurodevelopmental traits were assessed using the Autism-Tics, ADHD, and Other Comorbidities (A-TAC) inventory, a structured parent-report interview designed for large-scale epidemiological studies.^25^ The A-TAC consists of 96 symptom-based items grouped into modules corresponding to diagnostic domains and has demonstrated good validity for many domains.^26,27^ In the present study, continuous symptom scores at age 9 or 12 years were used. Domains examined included autism (total score and subdomains of social communication problems and restricted/repetitive behaviors and interests [RRBI]), ADHD (total score and subdomains of inattention and impulsivity/hyperactivity), learning disorders, tic disorders, ODD, conduct disorder, OCD, sensory perception problems (e.g., oversensitive to touch or sounds/noise), and sleep problems. A-TAC items are scored on a three-point scale (0 = no, 0.5 = yes, to some extent, 1 = yes) and domain scores represent the sum of item responses. Raw scores were used for descriptive analyses for interpretability, whereas standardized scores were used in the twin analyses for comparison across domains.

### Statistical Analysis

All analyses were conducted in R.^28^ Twin modeling analyses were conducted using the OpenMx package in R.^29^

#### Sample characteristics and phenotypic correlation

Descriptive statistics for demographic characteristics and neurodevelopmental trait scores were calculated for individuals with and without the broad ARFID phenotype, with between-group differences evaluated using generalized estimating equations with robust sandwich standard errors to account for nonindependence of twins within pairs. Phenotypic correlations between the broad ARFID phenotype and continuous neurodevelopmental traits were estimated as liability-scale polyserial correlations using full-information maximum likelihood in OpenMx.

#### Twin correlations and assumptions

The classical twin design estimates genetic and environmental contributions to individual differences by comparing similarity between monozygotic (MZ) twins, who share 100% of their segregating alleles, and dizygotic (DZ) twins, who share on average 50%. Both twin types are assumed to share environmental influences to a similar extent. Within-trait cross-twin correlations were calculated separately for MZ and DZ twin pairs to provide initial indications of genetic and environmental influences on each phenotype. Cross-twin cross-trait correlations between the broad ARFID phenotype and neurodevelopmental traits were then estimated to assess the extent to which phenotypic associations reflected shared genetic or environmental factors. Additive genetic influences (A) are suggested when MZ correlations exceed DZ correlations; dominant genetic influences (D) are suggested when DZ correlations are less than half of MZ correlations; shared environmental influences (C) are suggested when DZ correlations exceed half of MZ correlations; and nonshared environmental influences (E) are indicated when MZ correlations are less than 1.0 for same-trait correlations or when MZ correlations are below the corresponding within-person phenotypic correlations for cross-twin cross-trait correlations. E also includes measurement error.

#### Twin modelling

Twin-model assumptions were evaluated by comparing increasingly constrained saturated models that tested equality of thresholds, means, and variances across twin order and zygosity within sex. Joint continuous-binary liability threshold models were fitted to decompose the variance of the broad ARFID phenotype and each neurodevelopmental trait, as well as their covariance, into additive genetic (A), dominant genetic (D), shared environmental (C), and nonshared environmental (E) components. Broad-sense heritability (H), representing total genetic influence, was calculated as the sum of additive and dominant genetic effects (H = A + D). In the classical twin design, C and D cannot be estimated simultaneously; therefore, separate ACE and ADE models were evaluated, and the AE model was examined as a nested submodel. Model fit was evaluated using likelihood-ratio tests comparing nested models, as well as Akaike’s Information Criterion (AIC), with lower values indicating better fit and parsimony. The best-fitting model was selected based on a combination of statistical fit and parsimony. Estimates with 95% Wald confidence intervals were obtained for variance components of each phenotype and for genetic and environmental correlations between the broad ARFID phenotype and neurodevelopmental traits, including the additive (r_A_), dominant (r_D_), shared environmental (r_C_), and nonshared environmental (r_E_) correlations, depending on the model. Broad genetic correlations (r_H_) were also calculated when ADE models were selected. To quantify the sources of phenotypic associations, the phenotypic correlation between the broad ARFID phenotype and each neurodevelopmental trait was decomposed into the proportion attributable to genetic and environmental components.

## Results

### Sample characteristics

The analytic sample included 602 (2.0%) individuals with and 29,772 without the broad ARFID phenotype (**Table 1**). Individuals with the broad ARFID phenotype were more frequently male (60.8%) than those without it (50.5%) and had significantly higher mean symptom scores across all neurodevelopmental traits.

**Table 1.** Characteristics of the study sample by broad ARFID phenotype status.

|  | Theoretical range | Broad ARFID phenotype<br>(n=602)<br>No. (%) |  | No broad ARFID phenotype<br>(n=29,772)<br>No. (%) |  | p<sup>a</sup> |
| --- | --- | --- | --- | --- | --- | --- |
| **Demographic characteristics** |  |  |  |  |  |  |
| Sex |  |  |  |  |  | <.001 |
| Male |  |  | 366 (60.8) |  | 15,035 (50.5) |  |
| Female |  |  | 236 (39.2) |  | 14,737 (49.5) |  |
| Zygosity |  |  |  |  |  | .04 |
| Monozygotic |  |  | 151 (25.1) |  | 8,959 (30.1) |  |
| Dizygotic – same sex |  |  | 230 (38.2) |  | 10,524 (35.4) |  |
| Dizygotic – opposite sex |  |  | 221 (36.7) |  | 10,289 (34.6) |  |
| Age at CATSS assessment |  |  |  |  |  | .004 |
| 9 years |  |  | 499 (82.9) |  | 23,477 (78.86) |  |
| 12 years |  |  | 103 (17.1) |  | 6,295 (21.14) |  |
| **Neurodevelopmental traits** |  | **n** | **Mean (SD)** | **n** | **Mean (SD)** |  |
| Autism – total score | 0-17 | 597 | 3.25 (3.61) | 29,689 | 0.79 (1.53) | <.001 |
| Autism – social communication problems | 0-12 | 602 | 2.09 (2.48) | 29,772 | 0.54 (1.08) | <.001 |
| Autism – RRBI | 0-5 | 599 | 1.17 (1.38) | 29,700 | 0.25 (0.61) | <.001 |
| ADHD – total score | 0-19 | 594 | 5.84 (5.33) | 29,666 | 2.03 (3.06) | <.001 |
| ADHD – inattention | 0-9 | 595 | 3.15 (2.92) | 29,685 | 1.05 (1.74) | <.001 |
| ADHD – impulsivity/hyperactivity | 0-10 | 597 | 2.69 (2.84) | 29,683 | 0.98 (1.66) | <.001 |
| Learning disorders | 0-3 | 602 | 0.80 (1.03) | 29,724 | 0.28 (0.61) | <.001 |
| Tic disorders | 0-3 | 600 | 0.49 (0.84) | 29,703 | 0.16 (0.45) | <.001 |
| ODD | 0-5 | 600 | 1.35 (1.43) | 29,696 | 0.45 (0.84) | <.001 |
| Conduct disorder | 0-5 | 599 | 0.36 (0.76) | 29,702 | 0.09 (0.35) | <.001 |
| OCD | 0-2 | 602 | 0.18 (0.45) | 29,722 | 0.04 (0.20) | <.001 |
| Perception problems | 0-4 | 602 | 0.74 (0.98) | 29,772 | 0.17 (0.46) | <.001 |
| Sleep problems | 0-3 | 602 | 0.35 (0.60) | 29,772 | 0.11 (0.33) | <.001 |

Abbreviations: ADHD: attention deficit hyperactivity disorder; ARFID: avoidant/restrictive food intake disorder; OCD: obsessive-compulsive disorder; ODD: oppositional defiant disorder; RRBI: restricted/repetitive behavior and interests; SD: standard deviation.
<sup>a</sup> P values were estimated using generalized estimating equations with robust sandwich standard errors to account for nonindependence of twins within pairs.

### Phenotypic correlations

The broad ARFID phenotype was positively correlated with all neurodevelopmental traits examined (**Table 2**). Phenotypic correlations ranged from 0.18 to 0.36, with the strongest associations observed for autism total score (r_PH_ = 0.36), and the weakest correlations for OCD (r_PH_ = 0.18) and conduct disorder (r_PH_ = 0.19).

**Table 2.** Phenotypic correlation between the broad ARFID phenotype and neurodevelopmental traits.

| **Neurodevelopmental trait** | **$r_{PH}$ with ARFID (95% CI)** |
| --- | --- |
| Autism – total score | 0.36 (0.33, 0.38) |
| Autism – social communication problems | 0.33 (0.31, 0.36) |
| Autism – RRBI | 0.35 (0.32, 0.37) |
| ADHD – total score | 0.35 (0.33, 0.38) |
| ADHD – inattention | 0.35 (0.32, 0.38) |
| ADHD – impulsivity/hyperactivity | 0.30 (0.27, 0.33) |
| Learning disorders | 0.26 (0.23, 0.29) |
| Tic disorders | 0.20 (0.18, 0.23) |
| ODD | 0.32 (0.29, 0.34) |
| Conduct disorder | 0.19 (0.17, 0.22) |
| OCD | 0.18 (0.15, 0.21) |
| Perception problems | 0.31 (0.29, 0.34) |
| Sleep problems | 0.21 (0.18, 0.24) |

Abbreviations: ADHD: attention deficit hyperactivity disorder; ARFID: avoidant restrictive food intake disorder; CI: confidence interval; OCD: obsessive-compulsive disorder; ODD: oppositional defiant disorder; $r_{PH}$ : phenotypic correlation; RRBI: restricted/repetitive behavior and interests.

### Twin correlations

For the broad ARFID phenotype and all neurodevelopmental traits, within-trait DZ correlations were less than half of MZ correlations, suggesting additive genetic influences and possible dominance genetic effects on these phenotypes (**eTable 1**). Cross-twin cross-trait correlations between the broad ARFID phenotype and neurodevelopmental traits were also generally higher among MZ than DZ twins, supporting genetic contributions to their phenotypic associations. Most DZ cross-trait correlations were approximately half the corresponding MZ correlations, suggesting additive genetic influences, with no clear indication of shared environmental or dominant genetic effects.

### Twin assumption testing

Constraining thresholds and means to be equal across twin order within sex did not significantly worsen model fit, whereas additional constraints on variances and equality across zygosity groups showed evidence of poorer fit (**eTable 2**). However, the observed differences between Twin 1 and Twin 2 were small in magnitude (**eTable 3**), and the distributions of the neurodevelopmental trait scores were substantially skewed (**eTable 4; eFigures 1-13**), which may have contributed to statistically detectable differences. We proceeded with twin modeling under the assumptions of equal thresholds, means, and variances across twin order and zygosity within sex while acknowledging departures from these assumptions as a study limitation.

**Figure 1.**
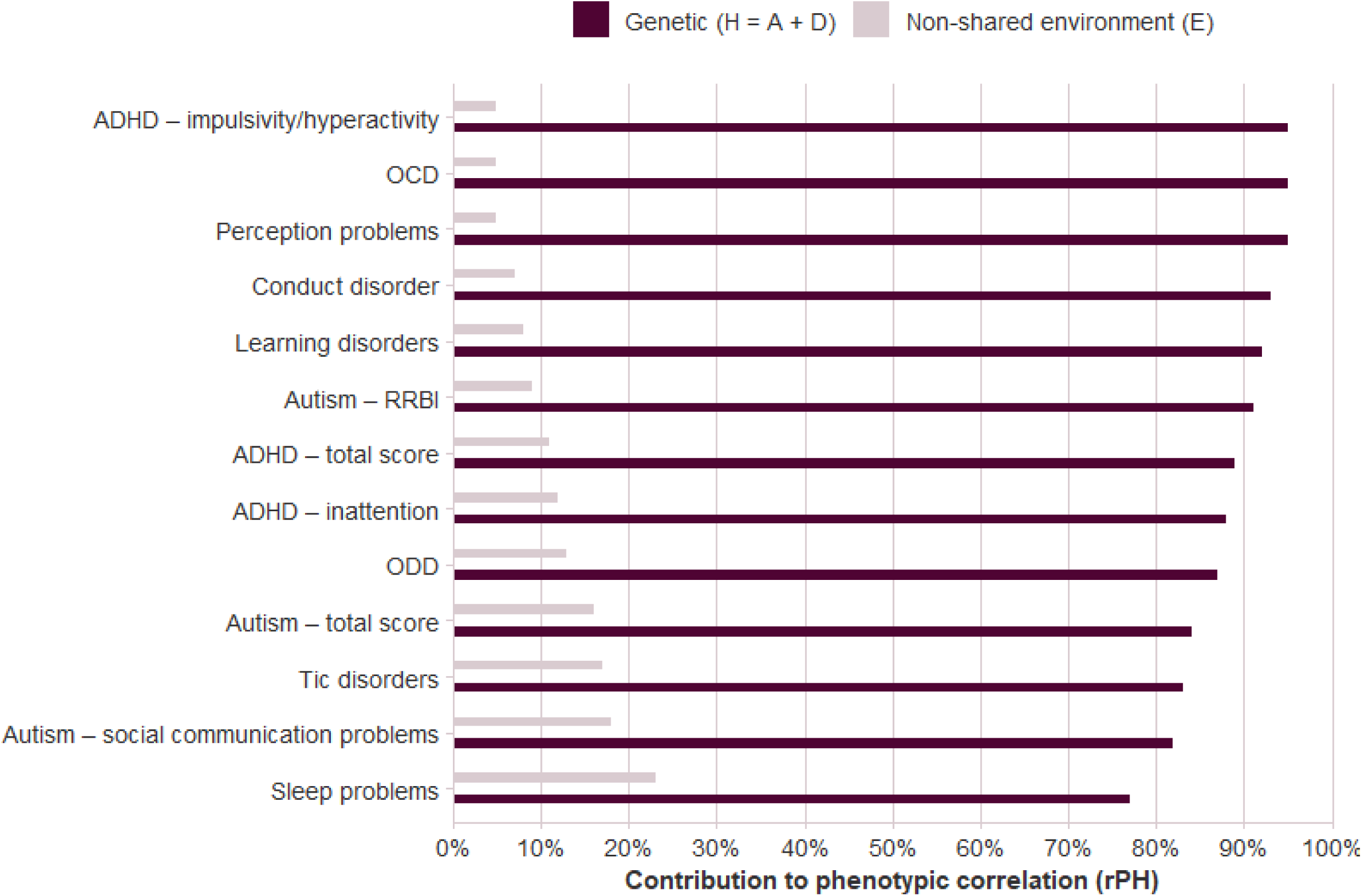
Proportion of contributions to phenotypic correlation (r_PH_) between the broad ARFID phenotype and neurodevelopmental traits by broad-sense genetic (H = A+D) and nonshared environmental (E) effects, ranked by the proportion attributable to H. The proportions attributable to H and E sum to 1 (100%) for each trait. Abbreviations: ADHD: attention deficit hyperactivity disorder; ARFID: avoidant restrictive food intake disorder; OCD: obsessive-compulsive disorder; ODD: oppositional defiant disorder; RRBI: restricted/repetitive behavior and interests.

### Bivariate twin models

ADE models provided the best fit for all associations (**Table 3**). Across bivariate models, broad-sense heritability estimates (H = A + D) ranged from 0.61 to 0.73 for the broad ARFID phenotype, and from 0.27 for OCD to 0.68 for autism total score among neurodevelopmental traits (**Table 4**). Broad genetic correlations between the broad ARFID phenotype and neurodevelopmental traits ranged from 0.27 for conduct disorder to 0.52 for autism-RRBI (**Table 4**). Nonshared environmental correlations were small and often nonsignificant. Decomposition of phenotypic correlations indicated that genetic influences accounted for most of the association between the broad ARFID phenotype and neurodevelopmental traits, explaining 77% to 95% of the phenotypic correlations, with the remaining variance attributable to nonshared environmental influences (**Figure 1; eTable 5**). Full results from ACE and AE models are presented in **eTables 6 and 7**.

**Table 3.** Model fit statistics for ACE, ADE, and AE models for associations between the broad ARFID phenotype and neurodevelopmental traits.

| Neurodevelopmental trait | Model | -2LL | Parameter | AIC | Comparison model | $\Delta\chi^2$ | $\Delta df$ | p |
| --- | --- | --- | --- | --- | --- | --- | --- | --- |
| Autism – total score | ACE | 85243.60 | 13 | 85269.60 | --- | --- | --- | --- |
|  | **ADE** | **85165.60** | **13** | **85191.60** | **ACE** | --- | --- | --- |
|  | AE<sup>a</sup> | 85243.60 | 10 | 85263.60 | ACE | 0 | 3 | 1 |
|  |  |  |  |  | ADE | 78.00 | 3 | <.001 |
| Autism – social communication problems | ACE | 86456.16 | 13 | 86482.16 | --- | --- | --- | --- |
|  | **ADE** | **86396.09** | **13** | **86422.09** | **ACE** | --- | --- | --- |
|  | AE | 86456.16 | 10 | 86476.16 | ACE | 0 | 3 | 1 |
|  |  |  |  |  | ADE | 60.07 | 3 | <.001 |
| Autism – RRBI | ACE | 86784.63 | 13 | 86810.63 | --- | --- | --- | --- |
|  | **ADE** | **86637.31** | **13** | **86663.31** | **ACE** | --- | --- | --- |
|  | AE | 86784.63 | 10 | 86804.63 | ACE | 0 | 3 | 1 |
|  |  |  |  |  | ADE | 147.31 | 3 | <.001 |
| ADHD – total score | ACE | 85850.59 | 13 | 85876.59 | --- | --- | --- | --- |
|  | **ADE** | **85742.21** | **13** | **85768.21** | **ACE** | --- | --- | --- |
|  | AE | 85850.59 | 10 | 85870.59 | ACE | 0 | 3 | 1 |
|  |  |  |  |  | ADE | 108.38 | 3 | <.001 |
| ADHD – inattention | ACE | 87465.32 | 13 | 87491.32 | --- | --- | --- | --- |
|  | **ADE** | **87275.50** | **13** | **87301.50** | **ACE** | --- | --- | --- |
|  | AE | 87465.32 | 10 | 87485.32 | ACE | 0 | 3 | 1 |
|  |  |  |  |  | ADE | 189.82 | 3 | <.001 |
| ADHD – impulsivity/hyperactivity | ACE | 86812.66 | 13 | 86838.66 | --- | --- | --- | --- |
|  | **ADE** | **86583.09** | **13** | **86609.09** | **ACE** | --- | --- | --- |
|  | AE | 86812.66 | 10 | 86832.66 | ACE | 0 | 3 | 1 |
|  |  |  |  |  | ADE | 229.57 | 3 | <.001 |
| Learning disorders | ACE | 88835.03 | 13 | 88861.03 | --- | --- | --- | --- |
|  | **ADE** | **88343.86** | **13** | **88369.86** | **ACE** | --- | --- | --- |
|  | AE | 88835.03 | 10 | 88855.03 | ACE | 0 | 3 | 1 |
|  |  |  |  |  | ADE | 491.17 | 3 | <.001 |
| Tic disorders | ACE | 87848.45 | 13 | 87874.45 | --- | --- | --- | --- |
|  | **ADE** | **87782.27** | **13** | **87808.27** | **ACE** | --- | --- | --- |
|  | AE | 88010.80 | 10 | 88030.80 | ACE | 162.35 | 3 | <.001 |
|  |  |  |  |  | ADE | 228.53 | 3 | <.001 |

**Table 3 (cont.).** Model fit statistics for ACE, ADE, and AE models for associations between the broad ARFID phenotype and neurodevelopmental traits.
| Neurodevelopmental trait | Model | -2LL | Parameter | AIC | Comparison model | $\Delta\chi^2$ | $\Delta df$ | p |
| --- | --- | --- | --- | --- | --- | --- | --- | --- |
| ODD | ACE | 87857.47 | 13 | 87883.47 | --- | --- | --- | --- |
|  | **ADE** | **87838.26** | **13** | **87864.26** | **ACE** | --- | --- | --- |
|  | AE | 87857.47 | 10 | 87877.47 | ACE | 0 | 3 | 1 |
|  |  |  |  |  | ADE | 19.22 | 3 | <.001 |
| Conduct disorder | ACE | 87135.30 | 13 | 87161.30 | --- | --- | --- | --- |
|  | **ADE** | **87107.74** | **13** | **87133.74** | **ACE** | --- | --- | --- |
|  | AE | 87135.30 | 10 | 87155.30 | ACE | 0 | 3 | 1 |
|  |  |  |  |  | ADE | 27.56 | 3 | <.001 |
| OCD | ACE | 90816.59 | 13 | 90842.59 | --- | --- | --- | --- |
|  | **ADE** | **90794.93** | **13** | **90820.93** | **ACE** | --- | --- | --- |
|  | AE | 90816.59 | 10 | 90836.59 | ACE | 0 | 3 | 1 |
|  |  |  |  |  | ADE | 21.66 | 3 | <.001 |
| Perception problems | ACE | 89537.48 | 13 | 89563.48 | --- | --- | --- | --- |
|  | **ADE** | **89438.78** | **13** | **89464.78** | **ACE** | --- | --- | --- |
|  | AE | 89802.67 | 10 | 89822.67 | ACE | 265.19 | 3 | <.001 |
|  |  |  |  |  | ADE | 363.89 | 3 | <.001 |
| Sleep problems | ACE | 90671.97 | 13 | 90697.97 | --- | --- | --- | --- |
|  | **ADE** | **90555.48** | **13** | **90581.48** | **ACE** | --- | --- | --- |
|  | AE | 90671.97 | 10 | 90691.97 | ACE | 0 | 3 | 1 |
|  |  |  |  |  | ADE | 116.50 | 3 | <.001 |

Abbreviations: -2LL: -2 log likelihood; $\Delta\chi^2$ : chi-square difference test; $\Delta df$ : difference in degree of freedom; A: additive genetic effects; ADHD: attention-deficit/hyperactivity disorder; AIC: Akaike's Information Criterion; ARFID: avoidant restrictive food intake disorder; C: shared environmental effects; D: dominance genetic effects; E: nonshared environmental effects; OCD: obsessive-compulsive disorder; ODD: oppositional defiant disorder; RRBI: restricted/repetitive behavior and interests;
<sup>a</sup>AE model is nested under ACE or ADE models. ACE and ADE are not nested models, so likelihood-ratio tests ( $\Delta\chi^2$ , $\Delta df$ , and p) are not reported for this comparison; model selection between ACE and ADE was based on AIC. The best-fitting models are **bolded**.

**Table 4.** Genetic and nonshared environmental variance as well as correlation estimates from the best-fitting (ADE) models for associations between the broad ARFID phenotype and neurodevelopmental traits.

| Neurodevelopmental trait | ARFID |  | Neurodevelopmental trait |  | Correlations |  |
| --- | --- | --- | --- | --- | --- | --- |
| | H | E | H | E | $r_H$ | $r_E$ |
| Autism – total score | 0.66 (0.54, 0.77) | 0.34 (0.23, 0.46) | 0.68 (0.67, 0.69) | 0.32 (0.31, 0.33) | 0.45 (0.39, 0.51) | 0.17 (0.08, 0.26) |
| Autism – social communication problems | 0.61 (0.50, 0.73) | 0.39 (0.27, 0.50) | 0.67 (0.65, 0.68) | 0.33 (0.32, 0.35) | 0.43 (0.36, 0.49) | 0.16 (0.08, 0.25) |
| Autism – RRBI | 0.66 (0.54, 0.77) | 0.34 (0.23, 0.46) | 0.57 (0.55, 0.59) | 0.43 (0.41, 0.45) | 0.52 (0.44, 0.60) | 0.08 (-0.01, 0.18) |
| ADHD – total score | 0.69 (0.58, 0.79) | 0.31 (0.21, 0.42) | 0.67 (0.65, 0.68) | 0.33 (0.32, 0.35) | 0.46 (0.40, 0.53) | 0.12 (0.02, 0.22) |
| ADHD – inattention | 0.68 (0.58, 0.79) | 0.32 (0.21, 0.42) | 0.57 (0.56, 0.59) | 0.43 (0.41, 0.44) | 0.49 (0.42, 0.57) | 0.11 (0.01, 0.22) |
| ADHD – impulsivity/hyperactivity | 0.67 (0.57, 0.76) | 0.33 (0.24, 0.43) | 0.67 (0.66, 0.69) | 0.33 (0.31, 0.34) | 0.43 (0.37, 0.49) | 0.05 (-0.05, 0.14) |
| Learning disorders | 0.71 (0.60, 0.81) | 0.29 (0.19, 0.40) | 0.67 (0.66, 0.69) | 0.33 (0.31, 0.34) | 0.35 (0.28, 0.41) | 0.07 (-0.04, 0.18) |
| Tic disorders | 0.69 (0.60, 0.79) | 0.31 (0.21, 0.40) | 0.40 (0.37, 0.42) | 0.60 (0.58, 0.63) | 0.33 (0.30, 0.35) | 0.08 (0.04, 0.12) |
| ODD | 0.71 (0.61, 0.80) | 0.29 (0.20, 0.39) | 0.58 (0.56, 0.60) | 0.42 (0.40, 0.44) | 0.43 (0.36, 0.50) | 0.12 (0.02, 0.22) |
| Conduct disorder | 0.73 (0.64, 0.82) | 0.27 (0.18, 0.36) | 0.60 (0.58, 0.61) | 0.40 (0.39, 0.42) | 0.27 (0.21, 0.33) | 0.04 (-0.06, 0.14) |
| OCD | 0.73 (0.64, 0.82) | 0.27 (0.18, 0.36) | 0.27 (0.24, 0.30) | 0.73 (0.07, 0.76) | 0.39 (0.29, 0.49) | 0.02 (-0.07, 0.11) |
| Perception problems | 0.65 (0.54, 0.77) | 0.35 (0.23, 0.46) | 0.52 (0.50, 0.54) | 0.48 (0.46, 0.50) | 0.51 (0.42, 0.59) | 0.04 (-0.06, 0.14) |
| Sleep problems | 0.73 (0.63, 0.82) | 0.27 (0.18, 0.37) | 0.42 (0.40, 0.45) | 0.58 (0.55, 0.60) | 0.29 (0.21, 0.38) | 0.12 (0.01, 0.23) |

Abbreviations: ADHD: attention deficit hyperactivity disorder; ARFID: avoidant restrictive food intake disorder; E: nonshared environmental effects; H (= A+D): broad-sense heritability representing total genetic effects, calculated as the sum of additive and dominant genetic effects; OCD: obsessive-compulsive disorder; ODD: oppositional defiant disorder; $r_E$ : correlation between ARFID and neurodevelopmental trait scores due to nonshared environmental influences; $r_H$ : correlation between ARFID and neurodevelopmental trait scores due to additive and dominance genetic influences; RRBI: restricted/repetitive behavior and interests.

## Discussion

In our study, the broad ARFID phenotype showed consistent positive associations with a broad range of neurodevelopmental traits, which were largely explained by shared genetic influences. These findings extend previous molecular genetic evidence. Bjorndal et al.^20^ reported a significant SNP-based genetic correlation between ARFID and ADHD (r_g_ = 0.21, SE = 0.10) but not autism and OCD, possibly reflecting limited statistical power due to small sample size and the restriction of SNP-based estimates to additive common variant effects. Koomar et al.^19^ reported a nominal association between ARFID total score assessed using the Nine-Item ARFID Screen^30^ and autism polygenic risk, with no association found with ADHD polygenic risk. In our study, positive genetic correlations were observed more consistently and broadly across multiple neurodevelopmental traits, which may reflect an advantage of the twin-based design that captures genetic influences beyond the additive effects of measured or imputed common variants indexed by SNP-based methods. Thus, prior molecular genetic studies may have underestimated the extent of genetic overlap between ARFID and neurodevelopmental traits. Importantly, genetic correlations indicate overlapping genetic influences that contribute to both phenotypes through potentially shared biological pathways, but these estimates do not directly evaluate causal relationships between neurodevelopmental traits and ARFID.^31^

The strongest phenotypic and genetic overlap was observed between the broad ARFID phenotype and autistic traits, particularly restricted/repetitive behaviors and interests, as well as perception problems and ADHD symptoms. Importantly, these associations are unlikely to be explained by item overlap, as none of the items included in these scales directly assessed eating-related behaviors. Although the phenotypic correlation with OCD was small (r_PH_ = 0.18), the genetic correlation was modest, and shared genetic factors accounted for almost all of this association. These findings suggest that genetic influences related to sensory processing, cognitive inflexibility, attentional regulation, and inhibitory control may also contribute to risk for avoidant/restrictive eating patterns.^32–34^ This pattern may be particularly relevant to two major clinical dimensions of ARFID: low appetite (i.e., lack of interest in food and eating) and sensory-based food avoidance (i.e., aversions to the taste, texture, or smell of foods).^35^ For example, genetic factors related to sensory processing and cognitive inflexibility may predispose individuals to avoidance based on the sensory properties of food and rigid food preferences, whereas genetic factors related to attentional regulation and inhibitory control may contribute to limited interest in eating or inconsistent intake. In contrast, the fear of aversive consequences dimension of ARFID may show less overlap with neurodevelopmental traits and may instead reflect anxiety-related processes or exposure to aversive eating-related experiences. Future research should examine whether genetic and environmental overlap with neurodevelopmental traits differs across ARFID dimensions. Together, these findings suggest that ARFID may represent a specific behavioral manifestation of broader neurodevelopmental traits when expressed in the domain of feeding and eating.

Environmental contributions to the covariance between the broad ARFID phenotype and neurodevelopmental traits were minimal. Nonshared environmental correlations were small and mostly nonsignificant, indicating that individual-specific experiences contribute little to why these traits co-occur, even though such factors may influence the development of each trait independently. Shared environmental influences (e.g., parents’ socioeconomic status) were not supported in model comparisons, as ADE models fit the data better than ACE models. However, because shared environmental and dominance genetic effects cannot be estimated simultaneously in the classical twin design, modest shared environmental influences cannot be ruled out entirely. These findings suggest that potential environmental risk factors for ARFID, such as aversive early feeding experiences and food-related events (e.g., choking, allergic reactions) or pre-existing medical conditions (e.g., gastrointestinal conditions),^36–39^ are likely to be disorder-specific rather than shared with neurodevelopmental traits.

### Strengths and Limitations

This study used a large, population-based twin cohort. The nationwide design, together with parent-report data and linkage of national health registers, enabled assessment of the broad ARFID phenotype and a broad range of co-occurring neurodevelopmental traits within a single population, while the twin design allowed for a direct estimation of genetic and environmental contributions to their overlap.

Several limitations should be considered. First, parent-reported neurodevelopmental traits may not fully correspond to clinical diagnoses and may introduce reporting bias. This may be relevant to the observed differences between MZ and DZ same-sex groups, which could partly reflect twin contrast effects where parents perceive twins as more distinct from one another. Future work should improve measurement for neurodevelopmental traits by incorporating multiple sources such as parent or self-reports, clinician ratings, or register-based diagnoses. Second, small Twin 1–Twin 2 differences were observed in some same-sex groups, which may reflect chance imbalance in randomization or skewed distributions of continuous neurodevelopmental traits. Given the small effect sizes, these differences were unlikely to be substantively meaningful. Third, the classical twin design cannot distinguish the specific biological mechanisms underlying shared genetic effects. Molecular genetic approaches will be important for clarifying underlying mechanisms. Fourth, the sample only consisted of Swedish twins, which may limit generalizability to other populations or cultural contexts. Replication in more diverse and non-European ancestry samples is warranted. Lastly, despite the large sample, the low prevalence of the broad ARFID phenotype limited power for examining sex differences. Larger pooled datasets may be needed to address this gap.

## Conclusions

The broad ARFID phenotype showed substantial genetic overlap with a number of neurodevelopmental traits, suggesting that ARFID may be better understood within a broader neurodevelopmental framework. Clinically, the observed genetic overlap indicates that neurodevelopmental traits are relevant to heterogeneity in ARFID, particularly for presentations characterized by sensory sensitivity, rigidity, or attentional difficulties. Together with prior evidence of elevated neurodevelopmental traits among individuals with ARFID, our results support systematic screening for neurodevelopmental difficulties in children presenting with ARFID, as well as monitoring eating disturbances in children with neurodevelopmental conditions. Future research should investigate whether specific ARFID presentations (sensory-based avoidance, low interest in eating, concern of aversive consequences) differ in their genetic overlap with neurodevelopmental traits, leverage molecular genetic data to identify specific biological pathways, and evaluate these associations across diverse populations and developmental stages to improve generalizability.

## Supporting information

eTables 1-7

## Data Availability

The individual-level data used in this study cannot be made publicly available because of Swedish legal and ethical restrictions. Researchers may apply for access to the data through the Swedish Twin Registry, subject to applicable ethical approval and data-access requirements.

## Author Contributions

Dr Dinkler had full access to all of the data in the study and takes responsibility for the integrity of the data and the accuracy of the data analysis.

*Study concept and design*: Qi, Taylor, Dinkler.

*Acquisition, analysis, or interpretation of data*: Lichtenstein, Lundstrom, Larsson, Bulik, Kuja-Halkola, Taylor, Dinkler.

*Drafting of the manuscript*: Qi, Dinkler.

*Critical revision of the manuscript for important intellectual content*: All authors.

*Statistical analysis*: Qi, Dinkler.

*Obtained funding*: Bulik, Dinkler.

*Administrative, technical, or material support*: Kuja-Halkola, Taylor.

*Study supervision*: Dinkler.

## Additional Contributions

We gratefully acknowledge the contribution of the participants in the Child and Adolescent Twin Study in Sweden (CATSS) and their families. We acknowledge the Swedish Twin Registry for access to data. The Swedish Twin Registry is managed by Karolinska Institutet and receives funding through the Swedish Research Council under grant 2017-00641.

## Data Sharing Statement

### Data available

No

### Explanation for why data not available

We are unable to share data due to Swedish laws. Researchers wishing to access the data themselves can apply directly to the Swedish Twin Registry.

---

**eFigure 1.**
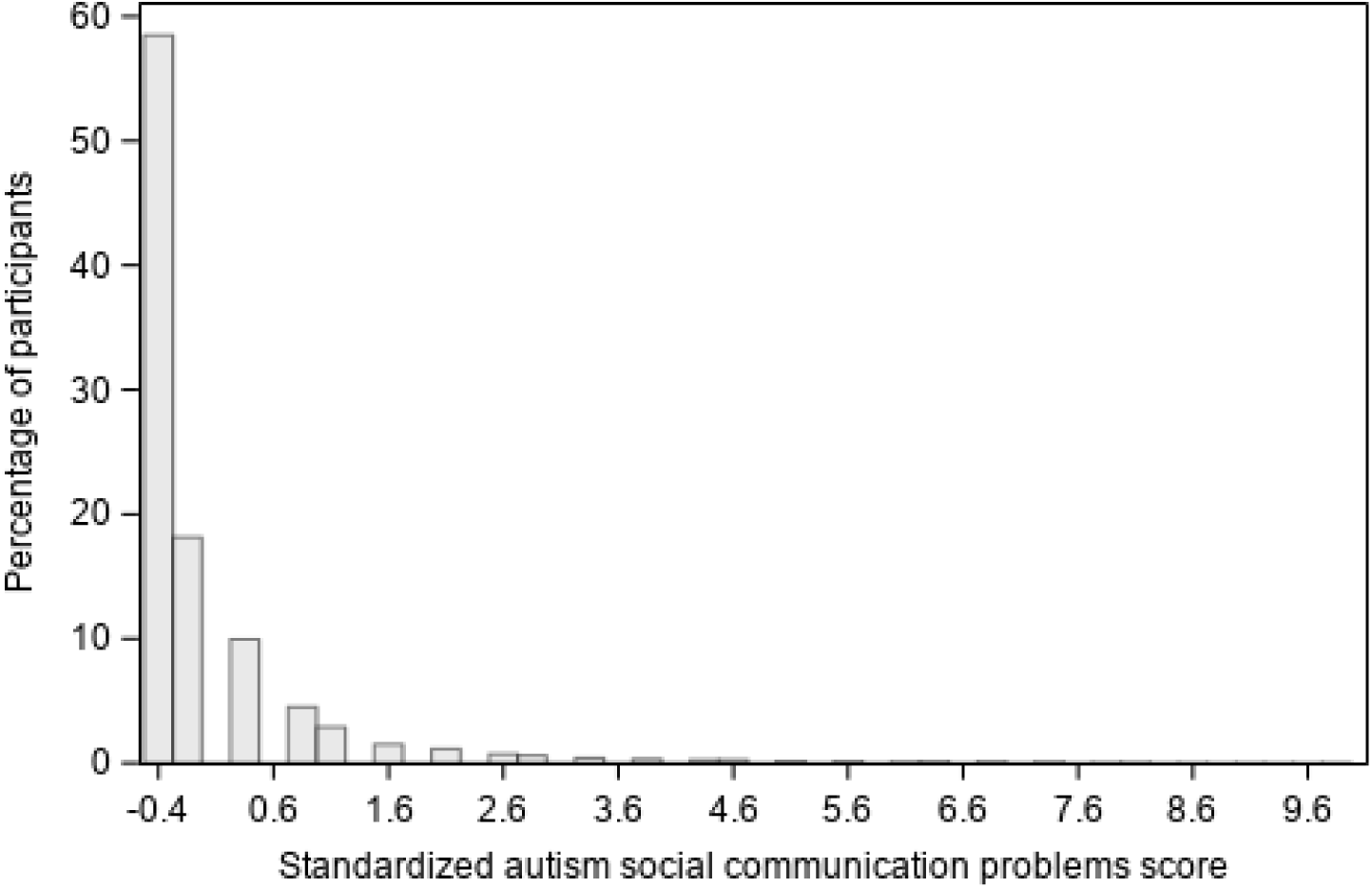
Distribution of standardized autism total score.

**eFigure 2.**
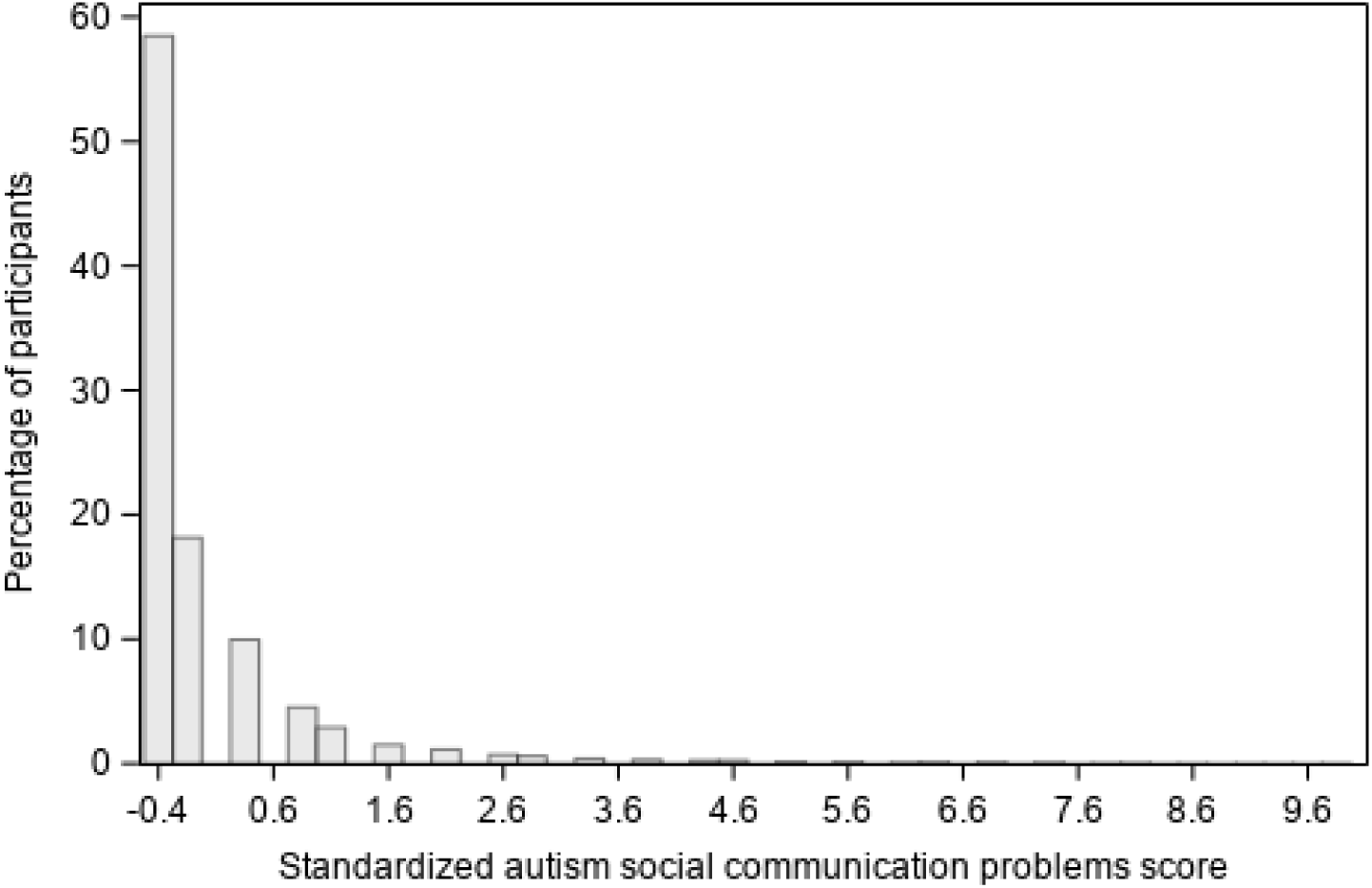
Distribution of standardized autism social communication problems score.

**eFigure 3.**
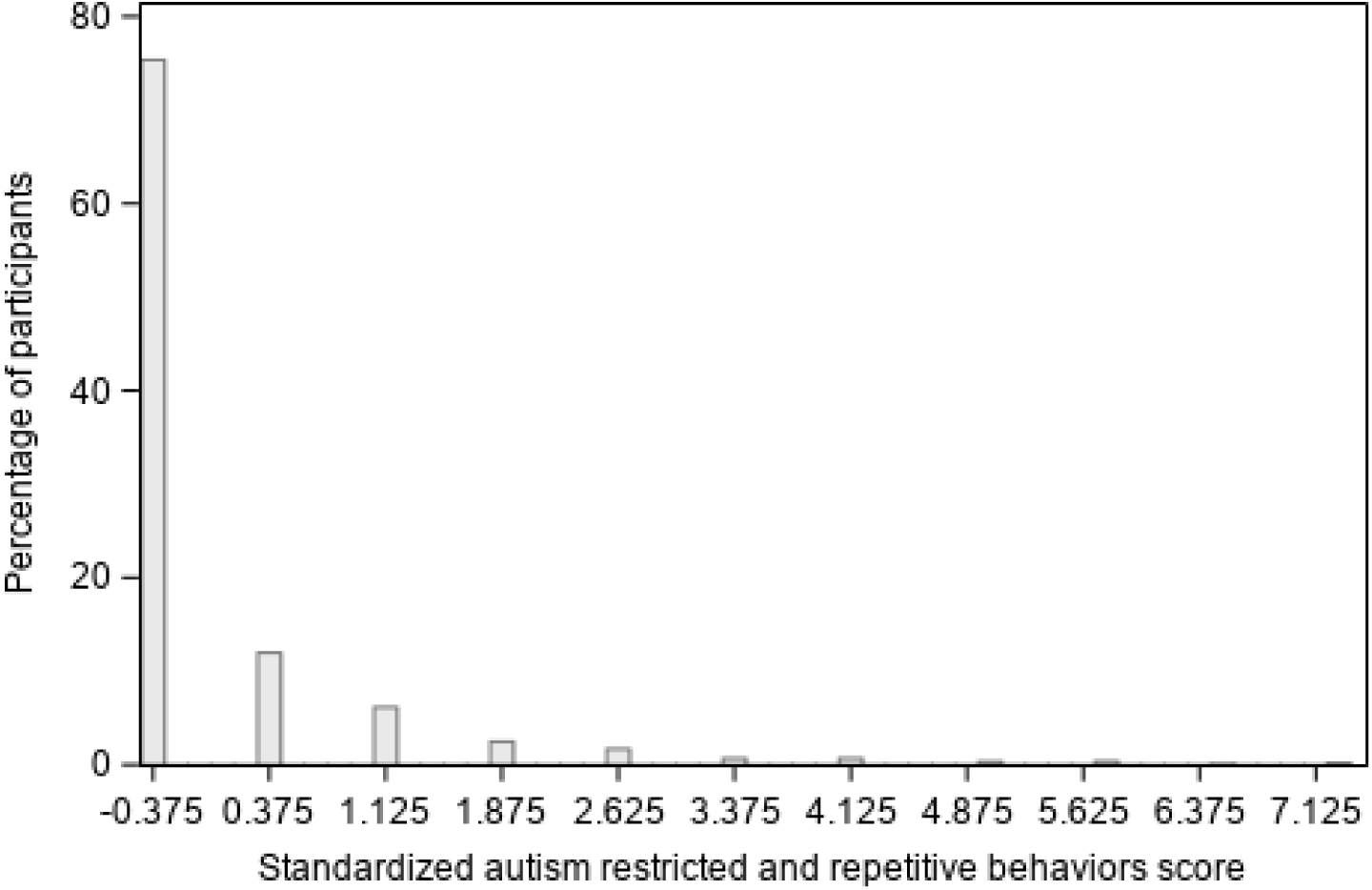
Distribution of standardized autism restricted and repetitive behaviors score.

**eFigure 4.**
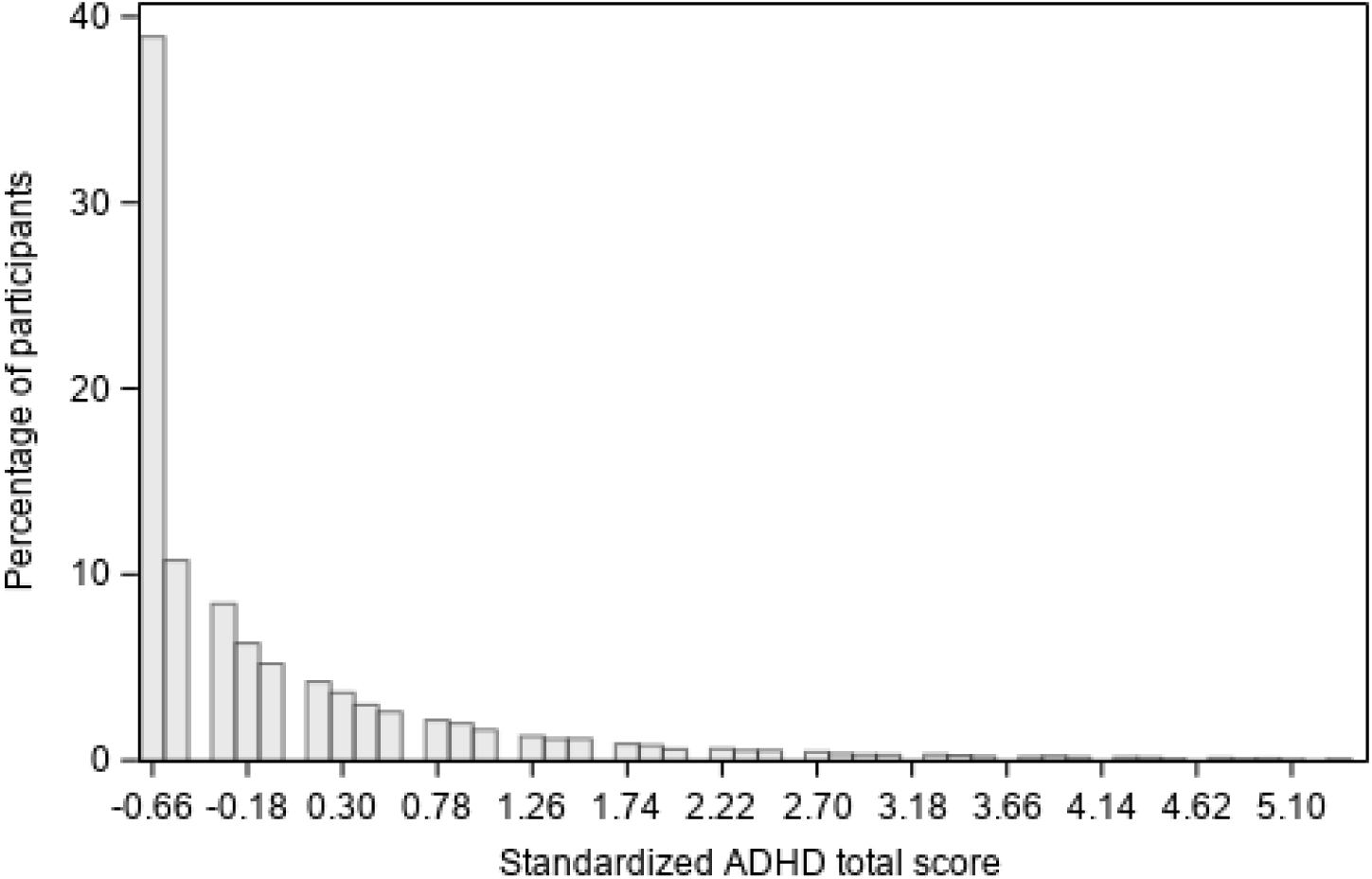
Distribution of standardized attention deficit hyperactivity disorder (ADHD) total score. inattention score.

**eFigure 5.**
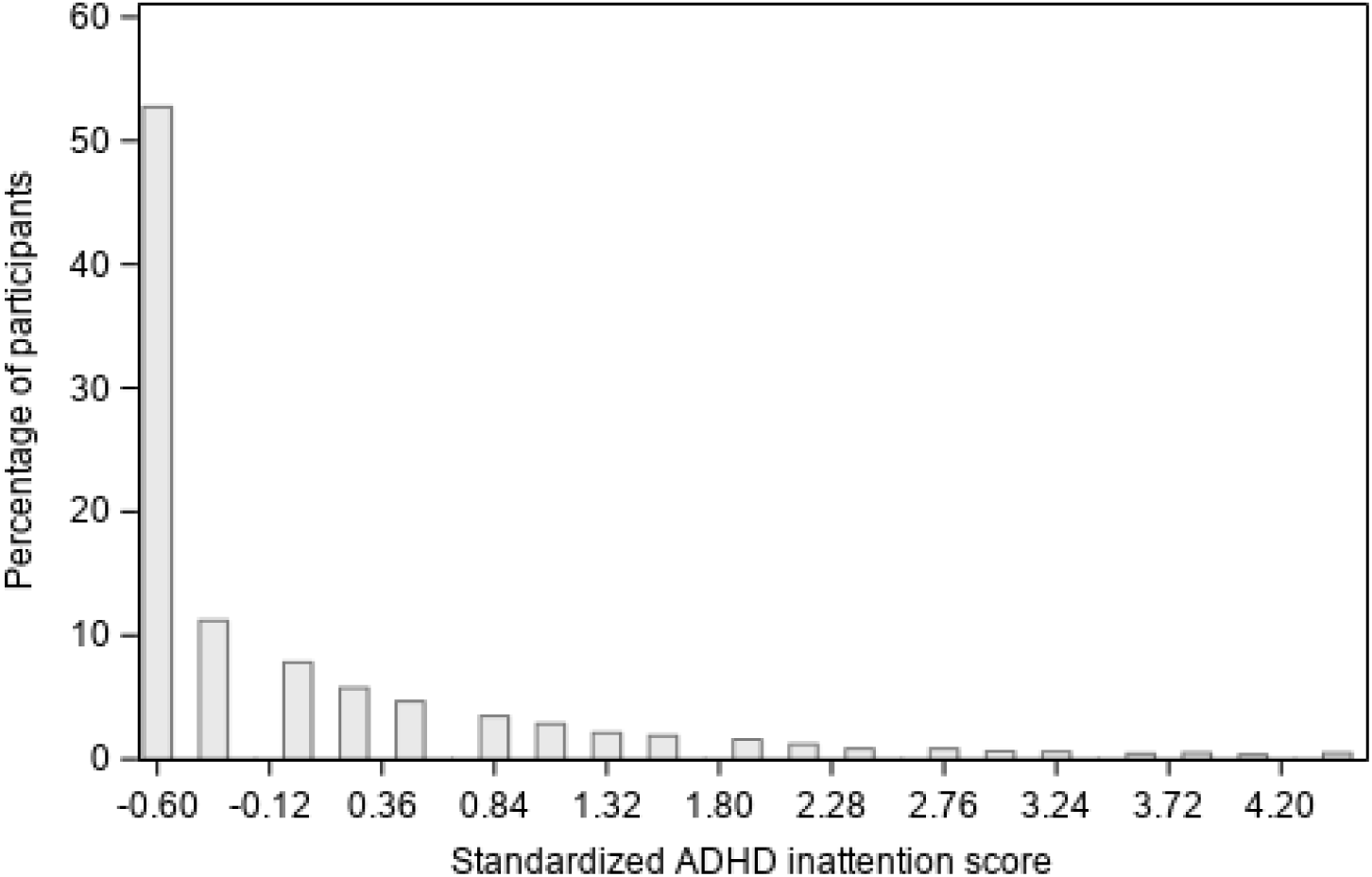
Distribution of standardized attention deficit hyperactivity disorder (ADHD) inattention score.

**eFigure 6.**
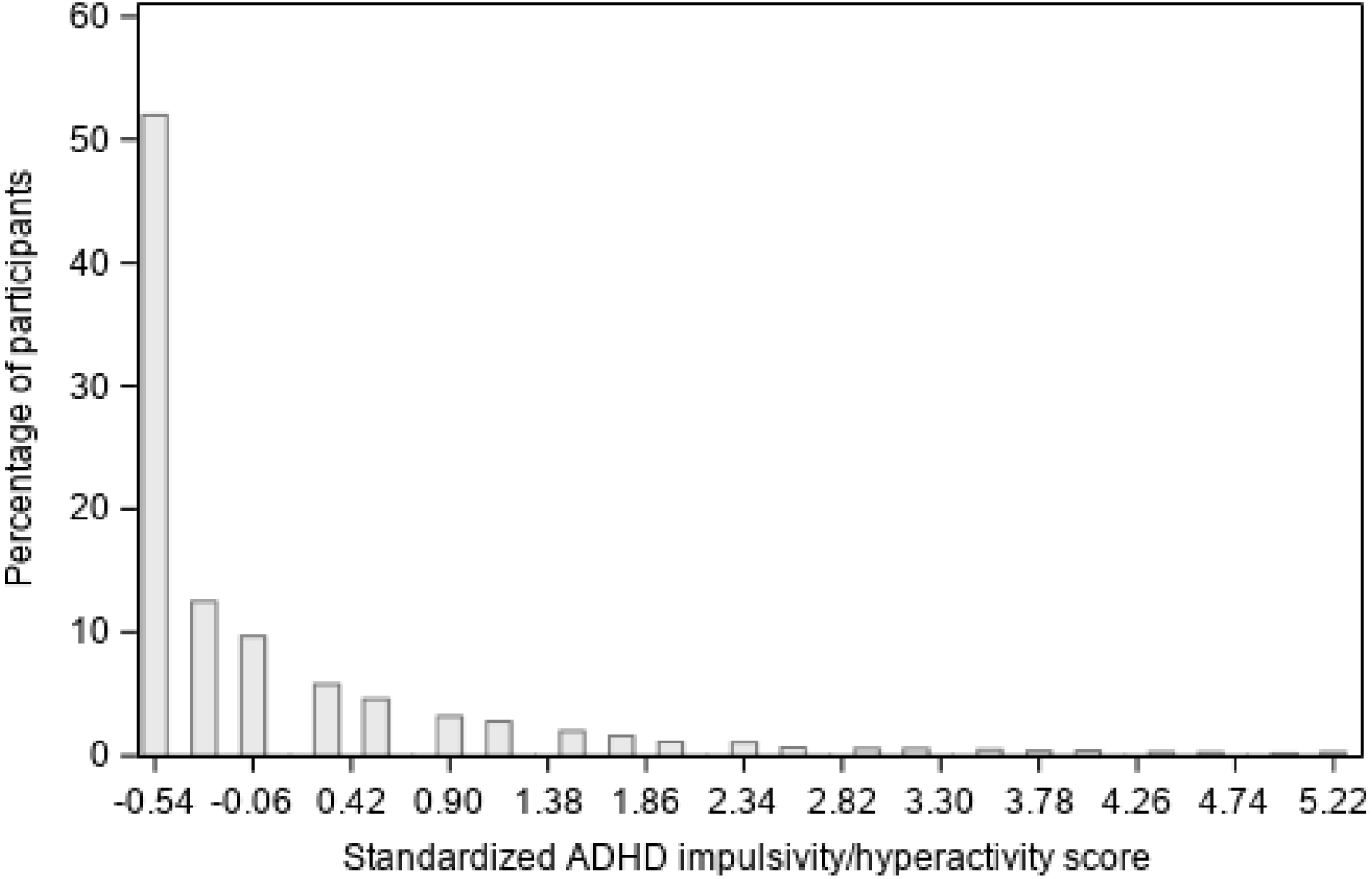
Distribution of standardized attention deficit hyperactivity disorder (ADHD) impulsivity/hyperactivity score.

**eFigure 7.**
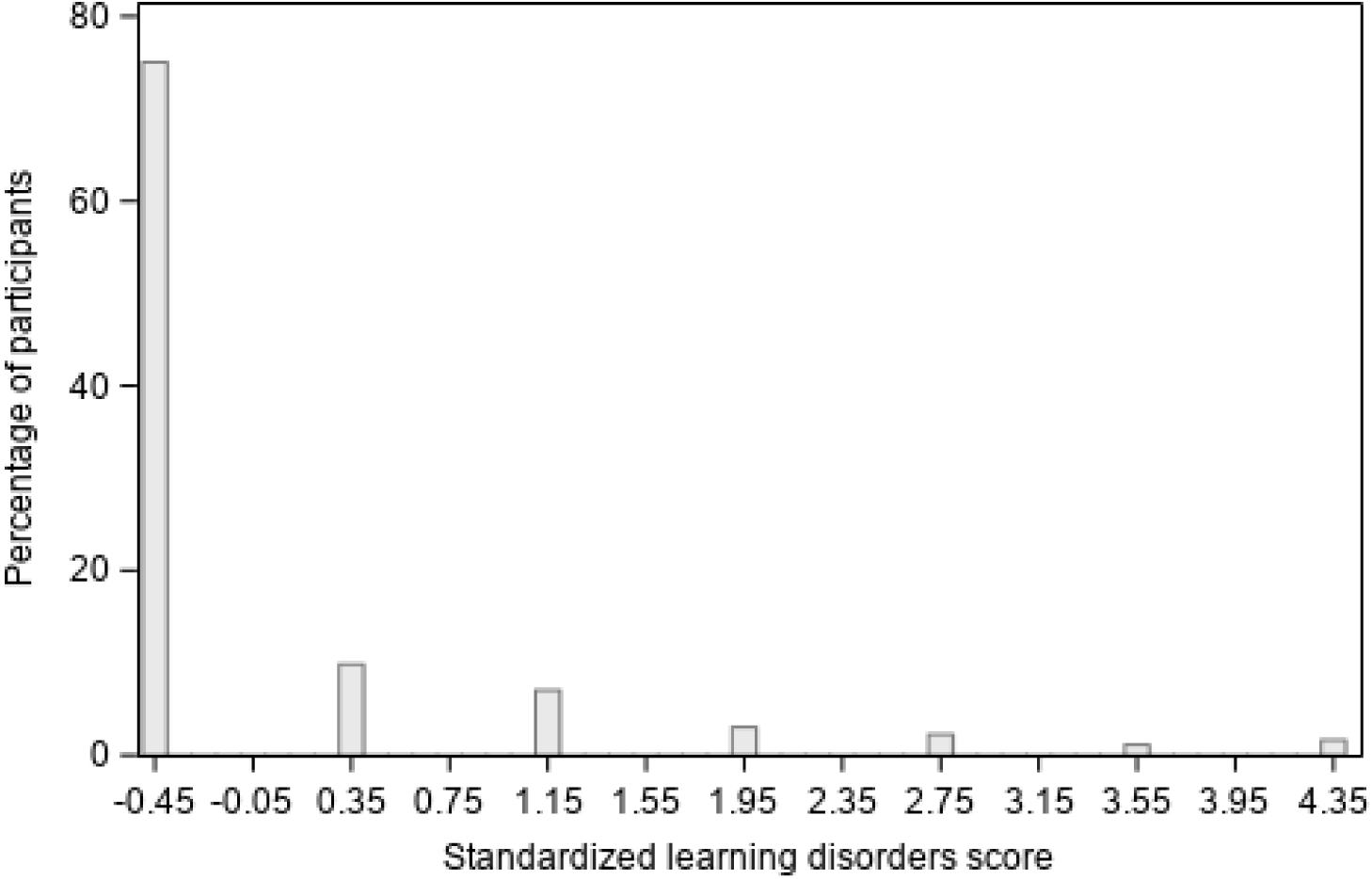
Distribution of standardized learning disorders score.

**eFigure 8.**
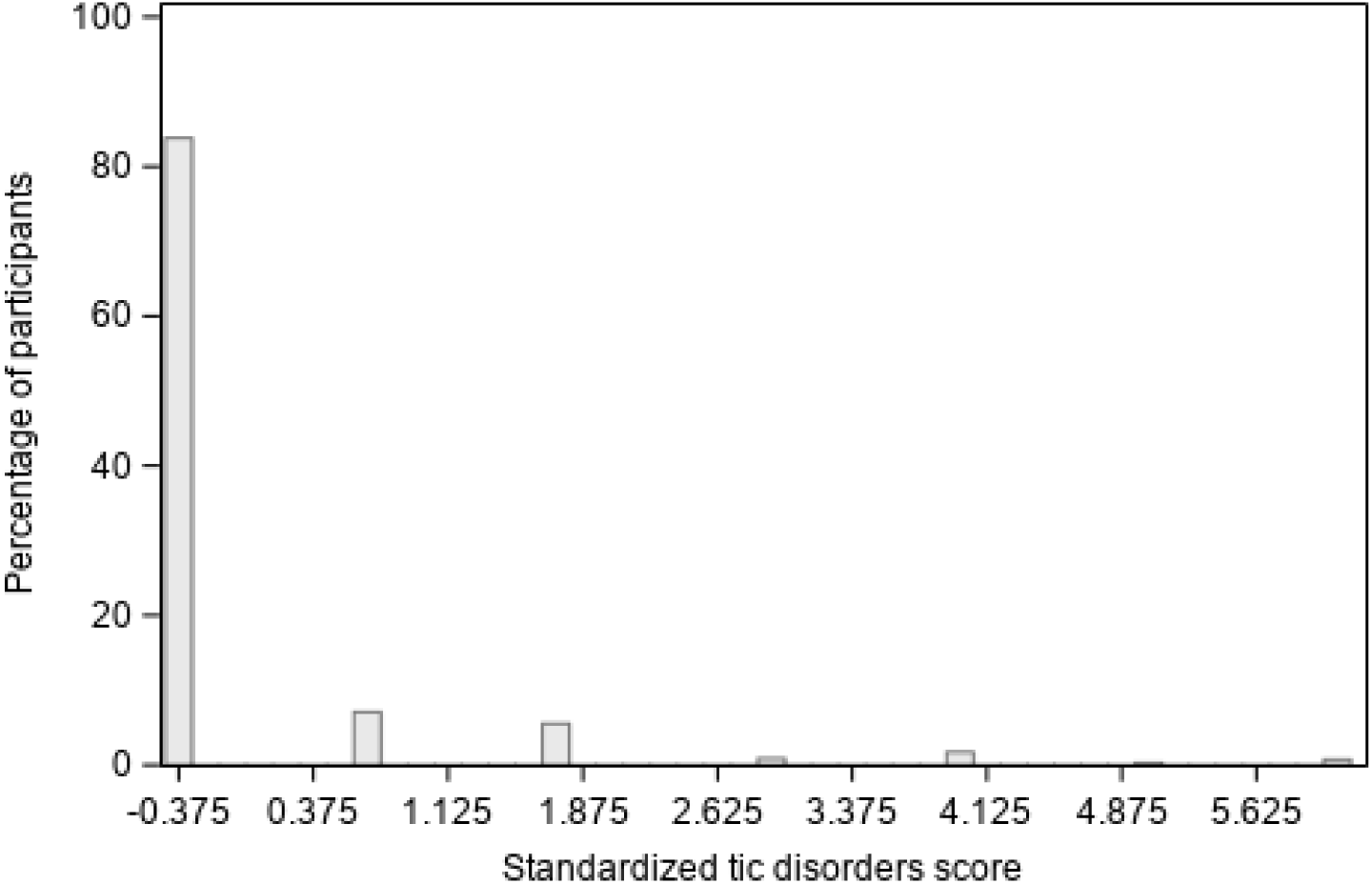
Distribution of standardized tic disorders score.

**eFigure 9.**
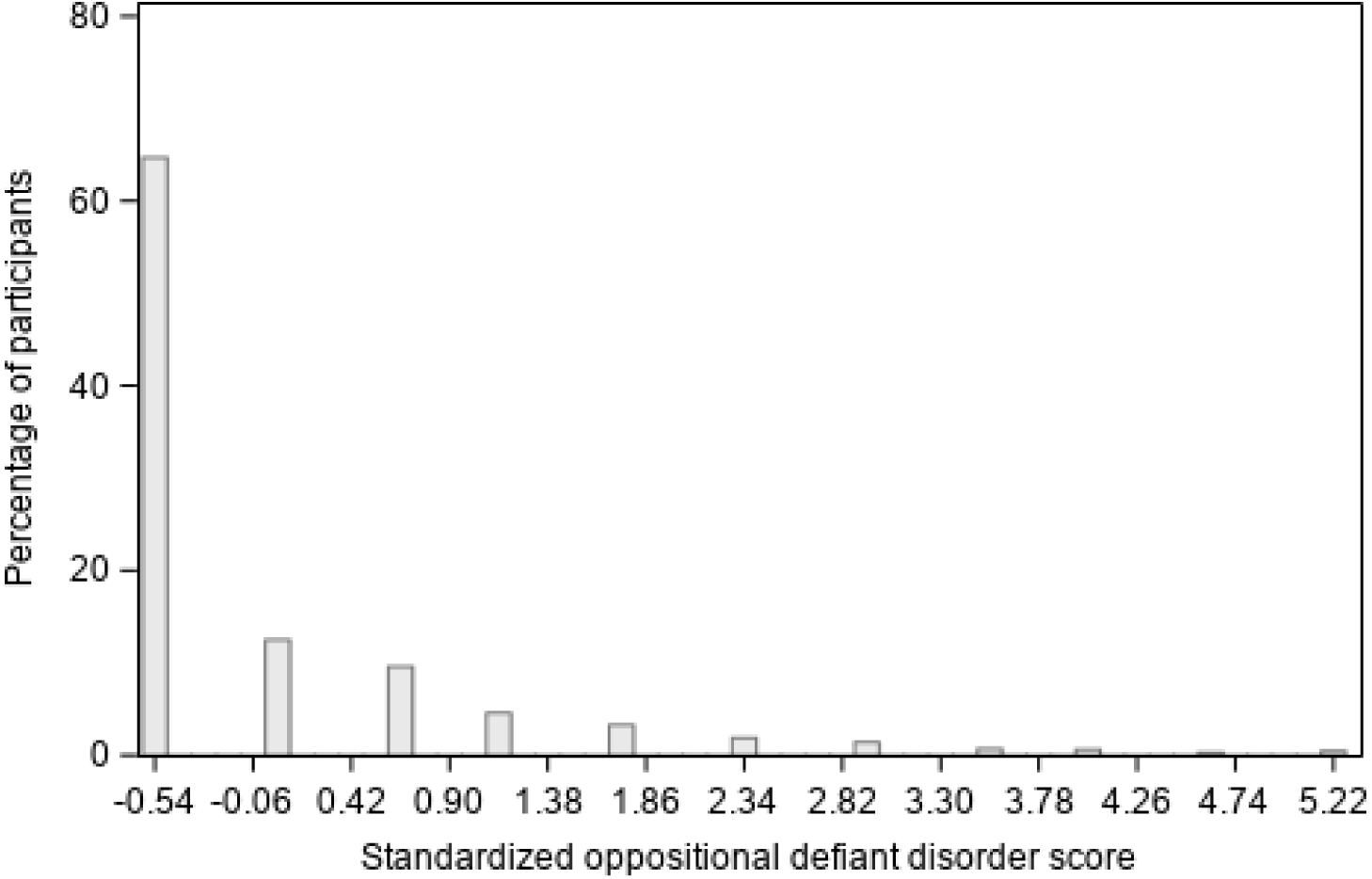
Distribution of standardized oppositional defiant disorder score.

**eFigure 10.**
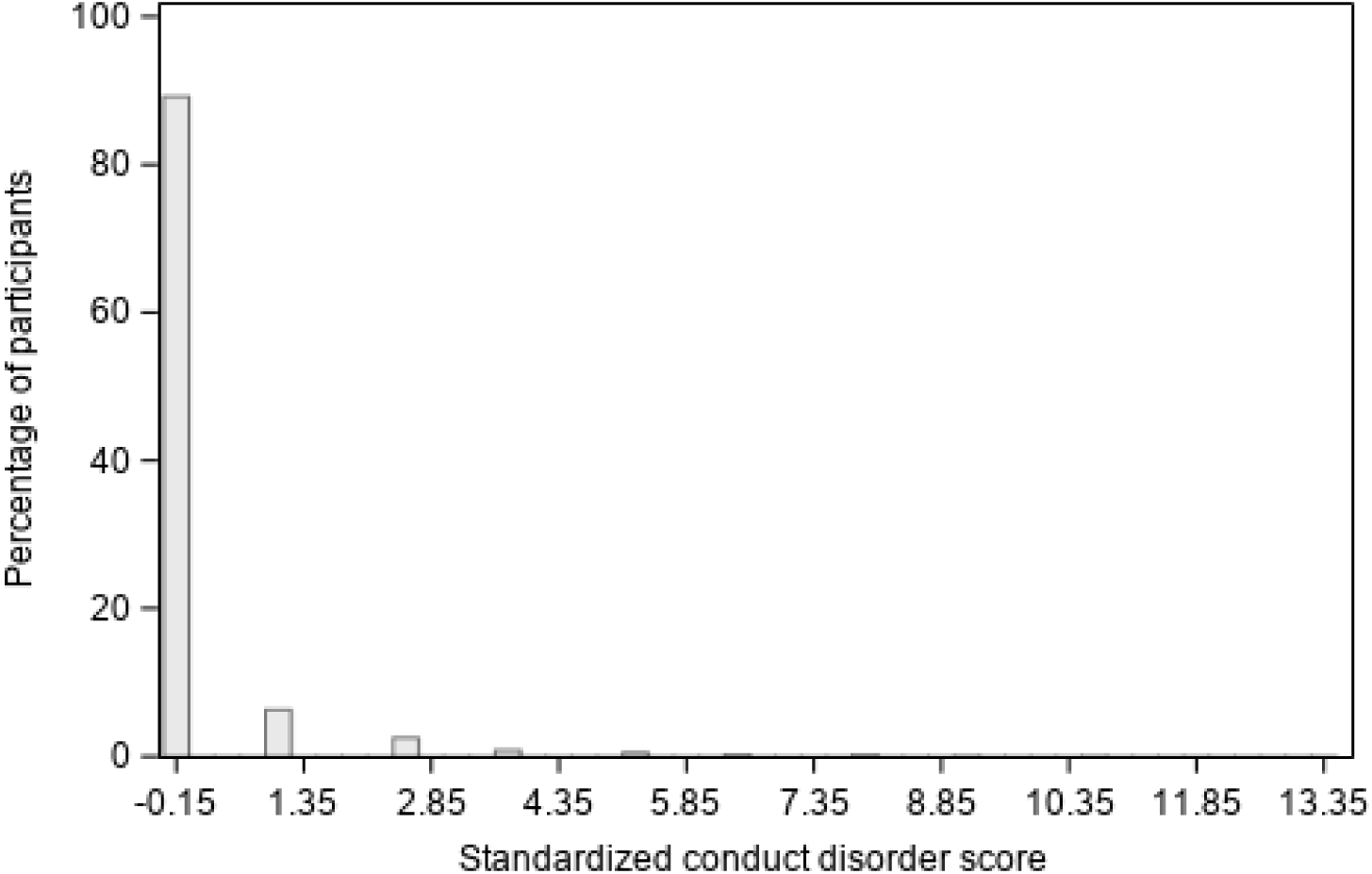
Distribution of standardized conduct disorder score.

**eFigure 11.**
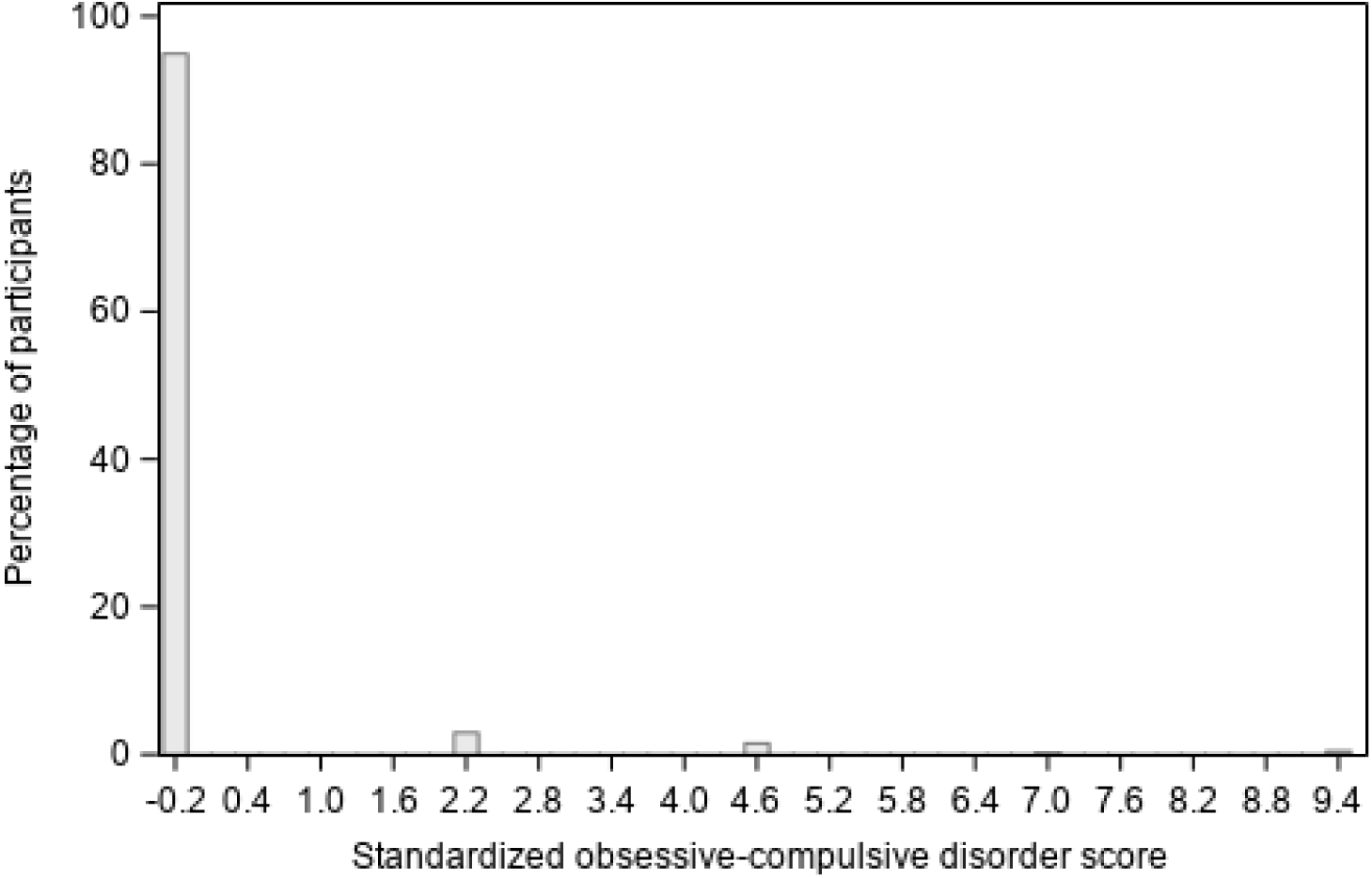
Distribution of standardized obsessive-compulsive disorder score.

**eFigure 12.**
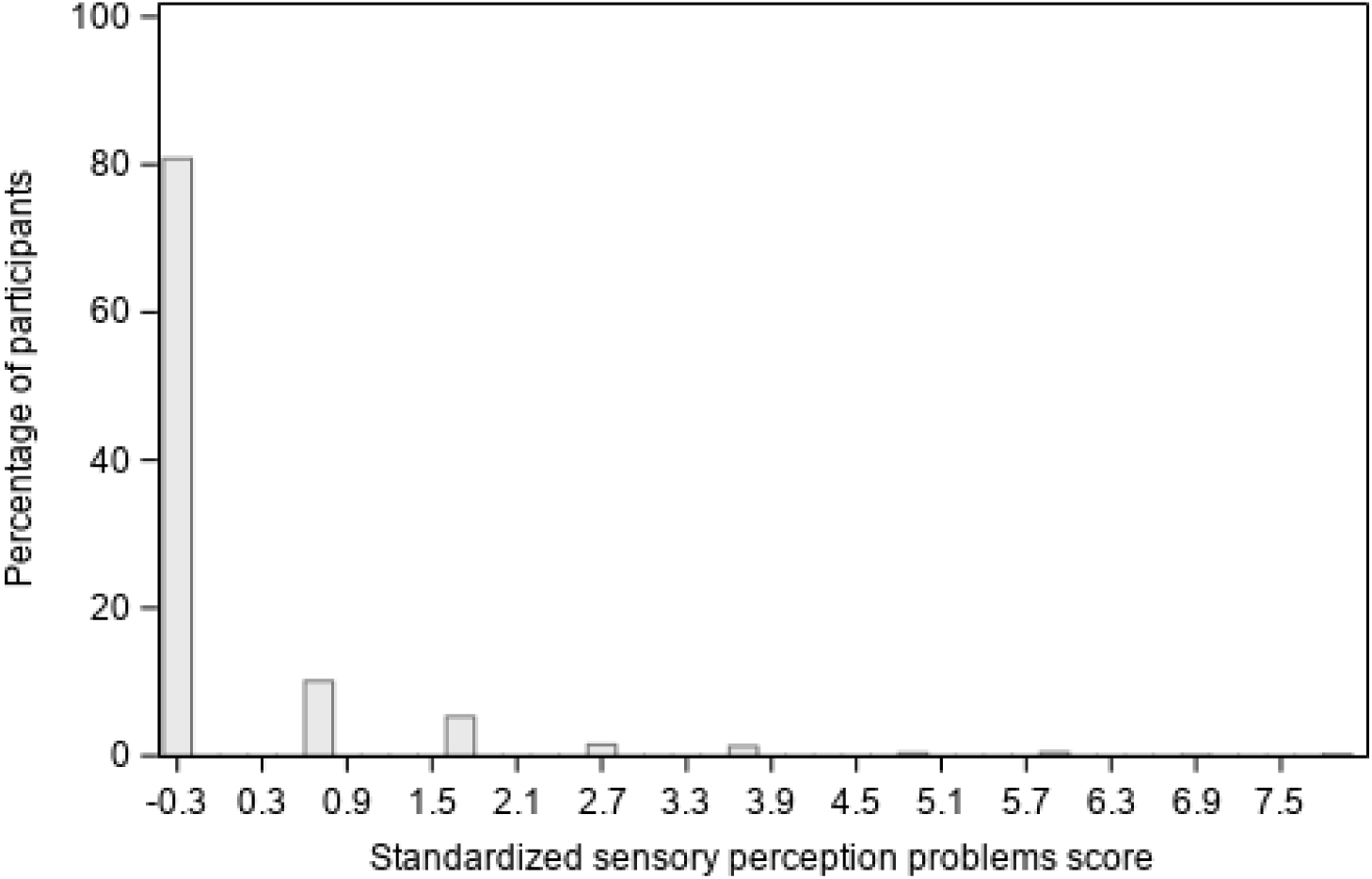
Distribution of standardized sensory perception problems score.

**eFigure 13.**
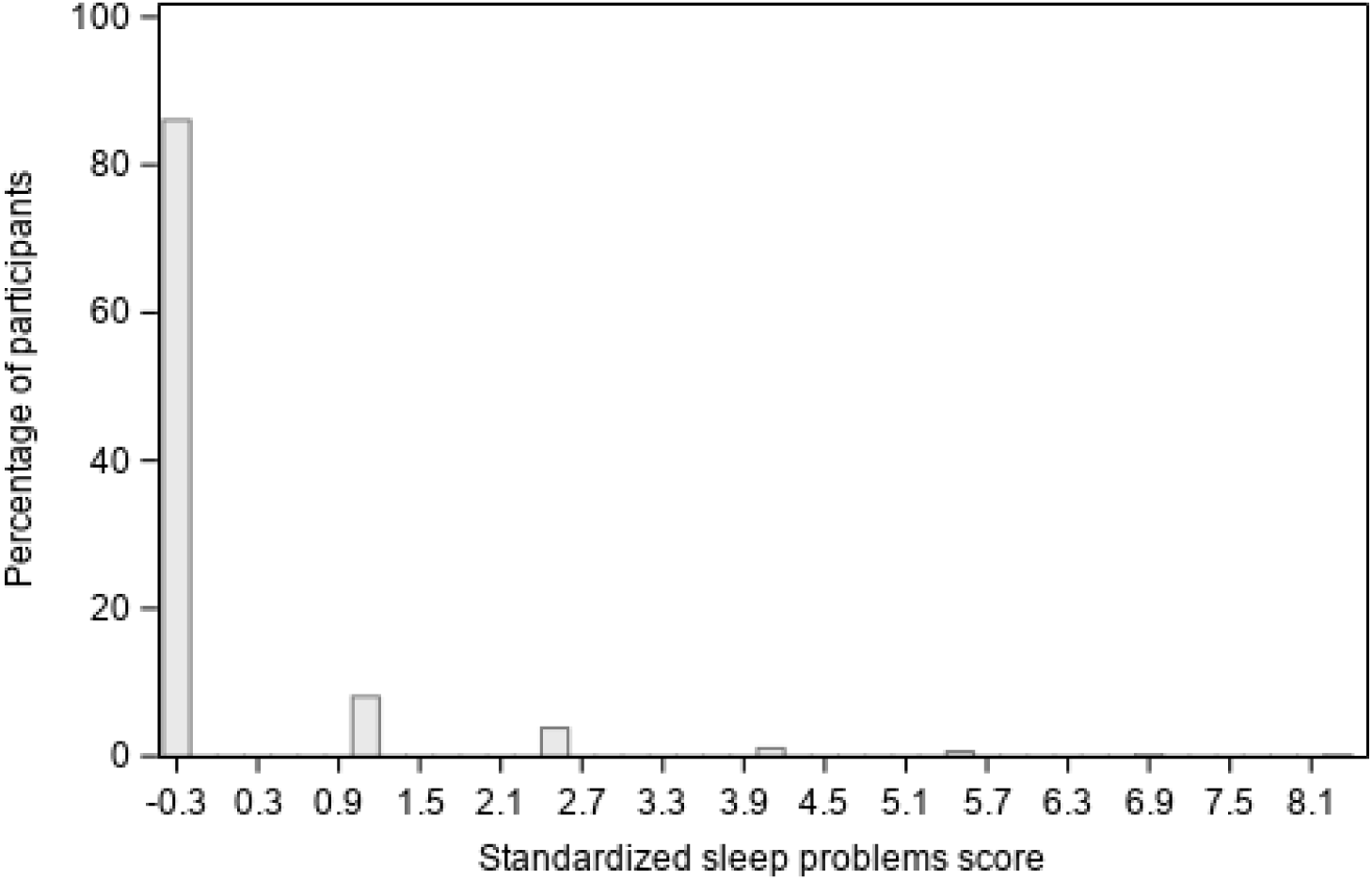
Distribution of standardized sleep problems score.

